# SDG5 ‘Gender Equality’ and the COVID-19 pandemic: a rapid assessment of health system responses in selected upper-middle and high-income countries

**DOI:** 10.1101/2022.09.09.22279765

**Authors:** Ellen Kuhlmann, Gabriela Lotta, Michelle Fernandez, Asha Herten-Crabb, Leonie Mac Fehr, Jaimie-Lee Maple, Ligia Paina, Clare Wenham, Karen Willis

## Abstract

The COVID-19 pandemic disrupted healthcare and societies, exacerbating existing inequalities for women and girls across every sphere. Our study explores health system responses to gender equality goals during the COVID-19 pandemic and inclusion in future policies. We apply a qualitative comparative approach, drawing on secondary sources and expert information; material was collected from March to July 2022. Australia, Brazil, Germany, the United Kingdom and USA were selected, reflecting upper-middle and high-income countries with established public health and gender policies but different types of healthcare systems and epidemiological and geo-political conditions. Three sub-goals of SDG 5 were analysed: maternity care and reproductive health, gender-based violence, and gender equality and women’s leadership. We found similar trends across countries. Pandemic policies strongly cut into women’s health, constrained prevention and support services and weakened reproductive rights, while essential maternity care services were kept open. Intersecting gender inequalities were reinforced, sexual violence increased and women’s leadership was weak. All healthcare systems failed to protect women’s health and essential public health targets. Yet there were relevant differences in the responses to increased violence and reproductive rights, ranging from some support measures in Australia to an abortion ban in the US. Our study highlights a need for revising pandemic policies through a feminist lens.

## Introduction

The COVID-19 pandemic ‘exacerbates existing inequalities for women and girls across every sphere – from health and the economy, to security and social protection’ (Feminist COVID-19 Collective, 2020, p.36). Health research illustrates the intersections between gender inequalities and racial, sexual, economic and other forms of social inequality during the pandemic (Adams-Prassel et al., 2020; Asefa et al, 2022; Bambra et al., 2021; Lalour et al., 2021; Lotta et al., 2021; Wenham et al., 2020). Lack of attention to women’s healthcare needs (of all ages, including girls) in pandemic policies was accompanied by antifeminist discourses and violation of reproductive rights. The recent US Supreme Court decision to no longer guarantee safe and legal access to abortion and related care (UN Women, 2022) is proof of these developments, but marks only the tip of the iceberg (Bojovic et al., 2021; UNFRA, 2020; Wenham et al., 2020).

An increase in gender inequalities during a major global public health crisis calls for a critical review of both pandemic policies and the United Nations Sustainable Development Goal 5 (SDG5) ‘Achieve gender equality and empower all women and girls’ (UN, 2020a) and its national/regional implementation, including gender mainstreaming approaches. However, a comprehensive gender-sensitive monitoring system of the impact of COVID-19 and pandemic policies is lacking; information is mainly collected by international NGOs (EuroHealthNet, 2021; IDLO, 2022; UN Women, 2020a,b,c) and feminist networks (Feminist Response to COVID-19 Collective, 2020; Morgan et al., 2022; Tomsick et al., 2022; Wenham et al., 2020).

Gender equality issues remain marginal (if not absent) in most high-level COVID-19 policy briefs and pandemic recovery plans and are often not included in key recommendations (Monti et al., 2021; OECD, 2020). Some statements mention women’s health and gender equality but lack systematic data and analysis (Unruh et al., 2022; WHO Euro, 2021). Against this backdrop, we sought to carry out a rapid assessment of the impact of COVID-19 on gender equality and the action taken in selected countries and areas of SDG5. Our research clarifies three major issues. How did the COVID-19 pandemic affect women’s health and gender equality goals in upper-middle and high income countries? What action (if any) was taken via by health policy to protect gender equality goals during the pandemic? What role do the SDG5 targets play in future policies and pandemic recovery plans?

## Methods

We apply a qualitative comparative approach based on explorative country case studies. The case studies draw on experts’ information and secondary sources, including published literature, websites and document analysis; material was collected in March/April 2022 with some amendments until July 2022. A rapid assessment and expert-based approach seem to be most helpful in a situation where information is scattered, research evidence poor and comparative data lacking.

### Connecting SDG5 ‘Gender Equality’ and SDG3 ‘Health’: an analytical framework

Our study is informed by health systems and governance theories and comparative health policy (Blank et al, 2018; Falkenbach et al., 2022; Gilson, 2012; Greer et al., 2022) and research into gender and health and feminist global health policy (Kuhlmann and Annandale, 2015; Lotta et al., 2021; UN Women, 2020a; Wenham, 2021, 2022). We focus on developments at the interface of SDG5 (UN, 2020a) and SDG3 ‘Ensure healthy lives and promote well-being for all at all ages’ (UN, 2020b). Inspired by the concept of ‘co-production’ (Falkenbach et al., 2022), we sought to identify intersecting targets. Four SDG5 sub-targets were selected:

- End all forms of discrimination against all women and girls everywhere (SDG 5.1).
- Eliminate all forms of violence against all women and girls in the public and private spheres (SDG5.2).
- Ensure women’s full and effective participation and equal opportunities for leadership at all levels of decision-making in political, economic and public life (SDG5.5).
- Ensure universal access to sexual and reproductive health and reproductive rights (SDG5.6) (https://www.undp.org/sustainable-development-goals#gender-equality).

For SDG3, we have chosen three sub-targets.

- Reduce maternal mortality (SDG3.1).
- Universal access to sexual and reproductive healthcare services (SDG3.7).
- Reduce violence everywhere (SDG target 16.1) (https://www.un.org/sustainabledevelopment/health).

Another important area includes women’s participation in new COVID-19 boards and female leadership in pandemic policy (Gabster et al., 2020). The Pan-European Commission on Health and Sustainable Development (2021) assessed the challenges posed by COVID-19 in the WHO European region and recommended, among others, improving gender equality and ‘explicit quotas…for the representation of women on public bodies that are involved in the formulation and implementation of health policy’ (Forman et al., 2022). The concept of ‘feminist global health security’ moves beyond quotas and connects intersecting inequalities to gender mainstreaming, using infectious disease prevention and pandemic policy as examples (Wenham, 2021).

Against this backdrop, we chose three interconnected topics of SDG3 and SDG5, considering different dimensions of gender equality.

1. Provision of maternity care and access to reproductive rights and services during the pandemic (SDG3.1, SDG3.7, SDG5.6).
2. Prevention of gender-based and sexual violence against women during the pandemic (SDG3 16.1, SDG5.2).
3. Support for gender equality and equity, including ensuring women’s participation in all areas and on all levels of health policy-making and strengthening female leadership (SDG 5.1, SDG5.5, and recent health policy debate and feminist concepts).

A generic conceptual framework (Figure 1) was developed, connecting the substance and the levels of analysis. ‘Substance’ focuses on the three selected thematic areas, while ‘levels’ are defined as follows: ‘*impact*’ of the COVID-19 pandemic on women’s health and service provision, ‘*action*’ taken on the institutional level (with some consideration of civil society engagement) and future ‘*policy*’ and pandemic recovery plans.

**Figure 1.**
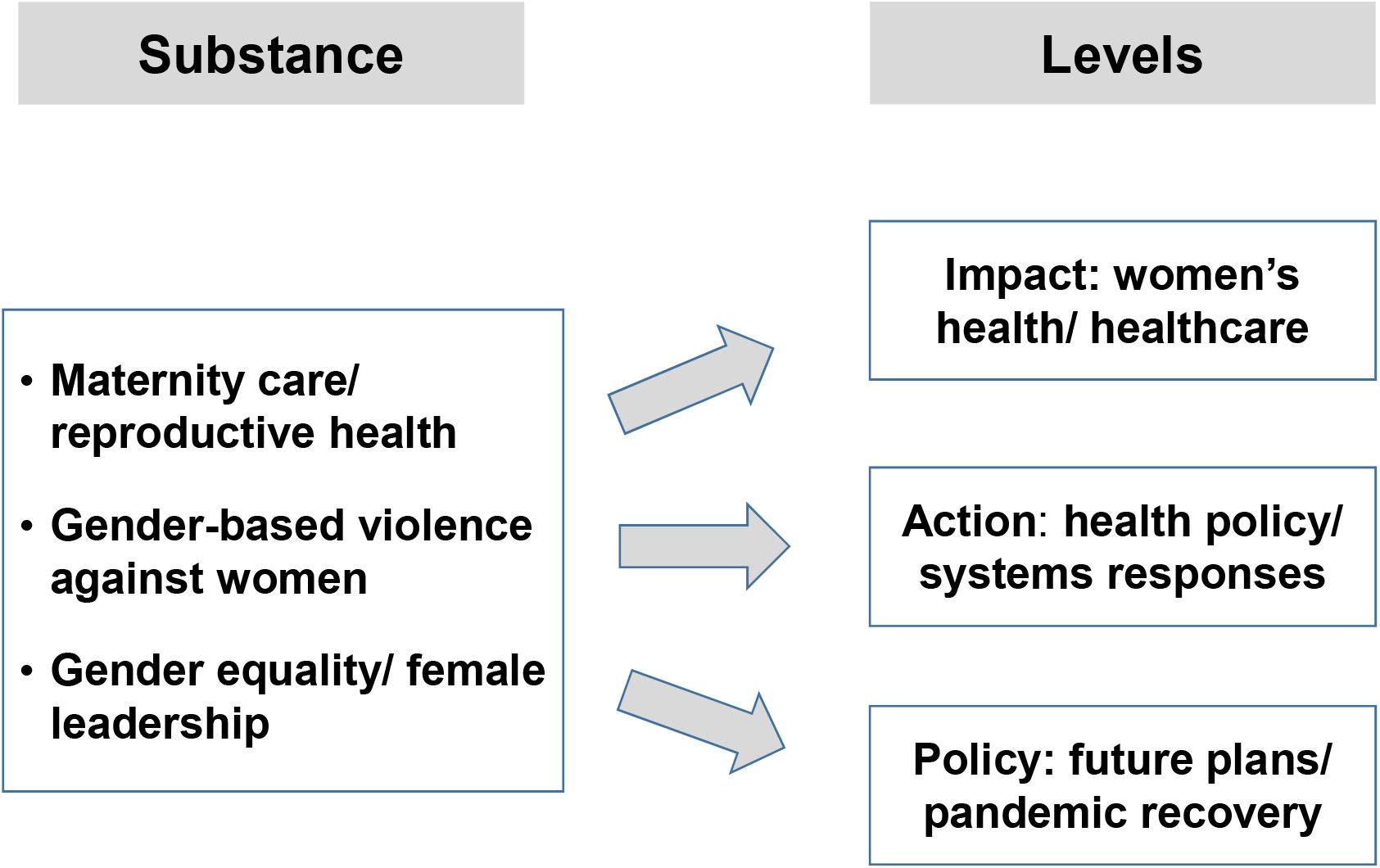
A generic assessment framework for SDG5 and COVID-19 policy Source: authors’ own figure

### Selecting the country cases: criteria and sample

Comparative health policy mainly draws on health system typologies (Burau et al., 2015), scores and quantitative indicators. COVID-19 revealed the weaknesses of these approaches. For instance, the global Epidemic Preparedness Index (EPI) put the US and UK in the highest and Brazil in the second-highest category of preparedness on a five-point score (Oppenheimer et al., 2019), but all three countries performed extremely poorly during COVID-19 (Table 1). System-based approaches largely ignore gender equality or limit the analysis to a few basic sex-based quantitative indicators. Feminist studies, in contrast, have developed alternative suggestions that take into account complexity and qualitative research (e.g., Morgan et al., 2022; Wenham et al., 2020; Wrede et al., 2006).

**Table 1.**
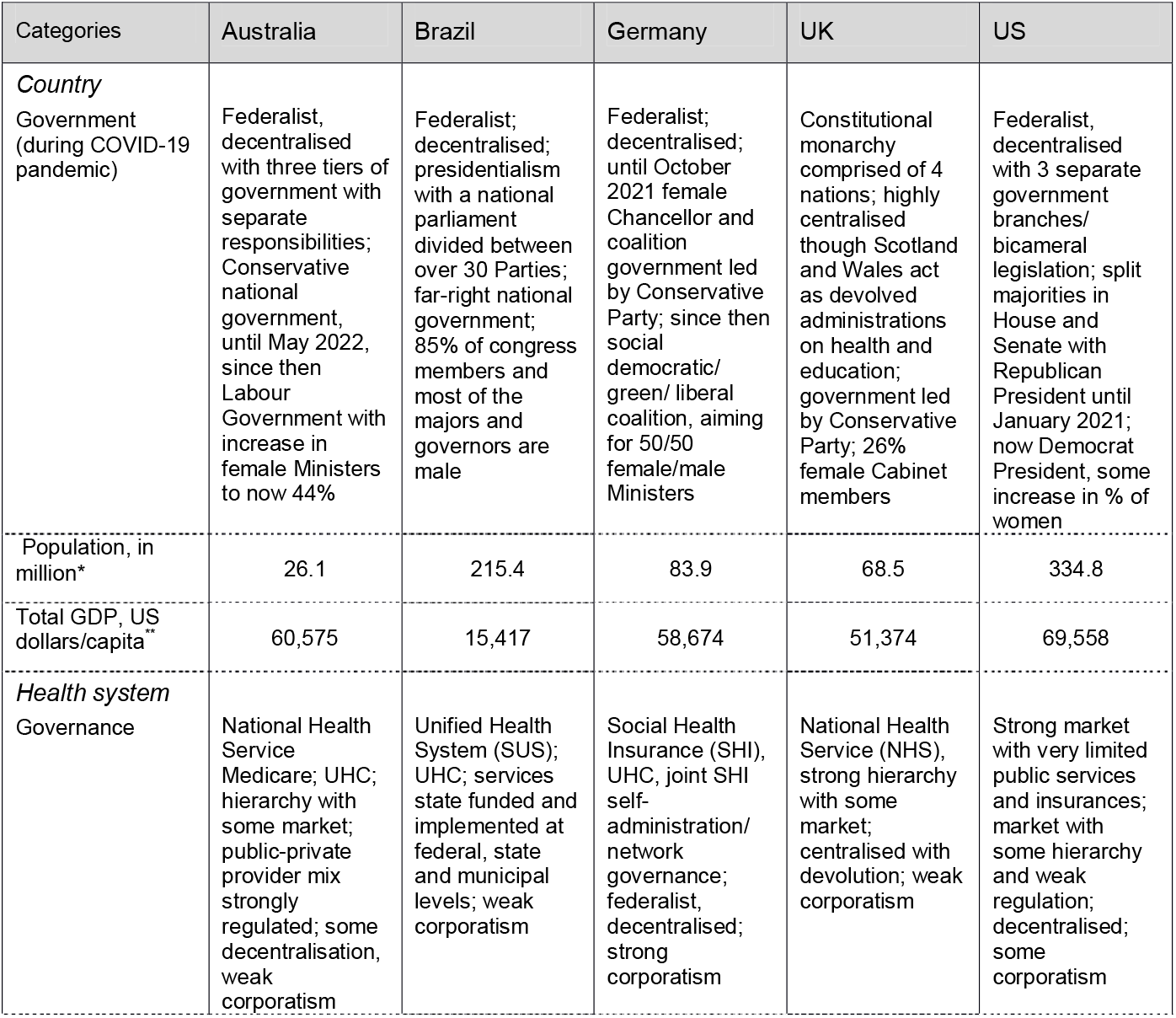
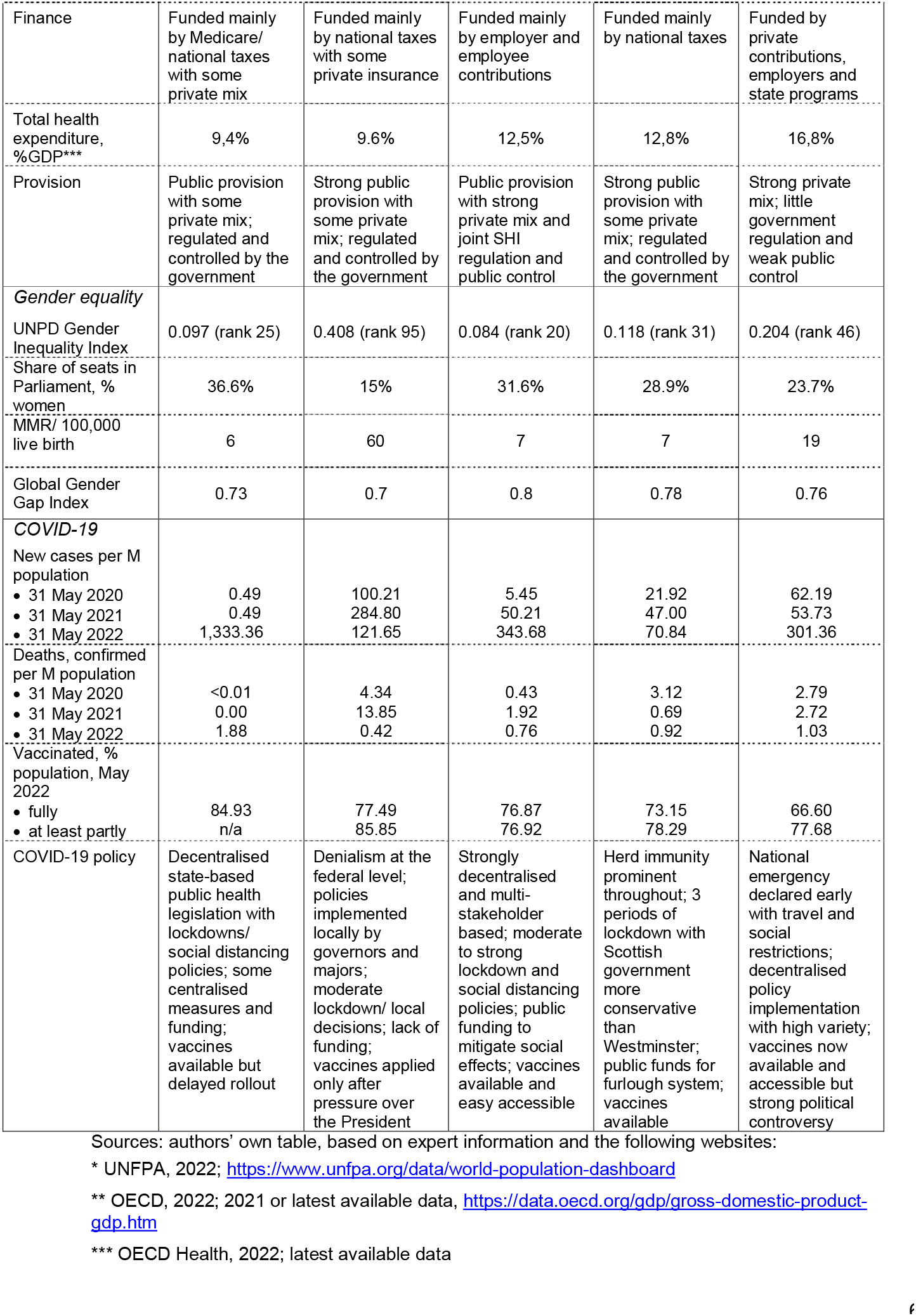
Mapping the country sample

| Categories | Australia | Brazil | Germany | UK | US |
| --- | --- | --- | --- | --- | --- |
| *Country*<br>Government<br>(during COVID-19<br>pandemic) | Federalist, decentralised with three tiers of government with separate responsibilities; Conservative national government, until May 2022, since then Labour Government with increase in female Ministers to now 44% | Federalist; decentralised; presidentialism with a national parliament divided between over 30 Parties; far-right national government; 85% of congress members and most of the majors and governors are male | Federalist; decentralised; until October 2021 female Chancellor and coalition government led by Conservative Party; since then social democratic/ green/ liberal coalition, aiming for 50/50 female/male Ministers | Constitutional monarchy comprised of 4 nations; highly centralised though Scotland and Wales act as devolved administrations on health and education; government led by Conservative Party; 26% female Cabinet members | Federalist, decentralised with 3 separate government branches/ bicameral legislation; split majorities in House and Senate with Republican President until January 2021; now Democrat President, some increase in % of women |
| Population, in million* | 26.1 | 215.4 | 83.9 | 68.5 | 334.8 |
| Total GDP, US dollars/capita** | 60,575 | 15,417 | 58,674 | 51,374 | 69,558 |
| *Health system*<br>Governance | National Health Service Medicare; UHC; hierarchy with some market; public-private provider mix strongly regulated; some decentralisation, weak corporatism | Unified Health System (SUS); UHC; services state funded and implemented at federal, state and municipal levels; weak corporatism | Social Health Insurance (SHI), UHC, joint SHI self-administration/ network governance; federalist, decentralised; strong corporatism | National Health Service (NHS), strong hierarchy with some market; centralised with devolution; weak corporatism | Strong market with very limited public services and insurances; market with some hierarchy and weak regulation; decentralised; some corporatism |

| Finance | Funded mainly by Medicare/ national taxes with some private mix | Funded mainly by national taxes with some private insurance | Funded mainly by employer and employee contributions | Funded mainly by national taxes | Funded by private contributions, employers and state programs |
| --- | --- | --- | --- | --- | --- |
| Total health expenditure, %GDP*** | 9,4% | 9.6% | 12,5% | 12,8% | 16,8% |
| Provision | Public provision with some private mix; regulated and controlled by the government | Strong public provision with some private mix; regulated and controlled by the government | Public provision with strong private mix and joint SHI regulation and public control | Strong public provision with some private mix; regulated and controlled by the government | Strong private mix; little government regulation and weak public control |
| *Gender equality* |  |  |  |  |  |
| UNPD Gender Inequality Index | 0.097 (rank 25) | 0.408 (rank 95) | 0.084 (rank 20) | 0.118 (rank 31) | 0.204 (rank 46) |
| Share of seats in Parliament, % women | 36.6% | 15% | 31.6% | 28.9% | 23.7% |
| MMR/ 100,000 live birth | 6 | 60 | 7 | 7 | 19 |
| Global Gender Gap Index | 0.73 | 0.7 | 0.8 | 0.78 | 0.76 |
| *COVID-19* |  |  |  |  |  |
| New cases per M population |  |  |  |  |  |
| • 31 May 2020 | 0.49 | 100.21 | 5.45 | 21.92 | 62.19 |
| • 31 May 2021 | 0.49 | 284.80 | 50.21 | 47.00 | 53.73 |
| • 31 May 2022 | 1,333.36 | 121.65 | 343.68 | 70.84 | 301.36 |
| Deaths, confirmed per M population |  |  |  |  |  |
| • 31 May 2020 | <0.01 | 4.34 | 0.43 | 3.12 | 2.79 |
| • 31 May 2021 | 0.00 | 13.85 | 1.92 | 0.69 | 2.72 |
| • 31 May 2022 | 1.88 | 0.42 | 0.76 | 0.92 | 1.03 |
| Vaccinated, % population, May 2022 |  |  |  |  |  |
| • fully | 84.93 | 77.49 | 76.87 | 73.15 | 66.60 |
| • at least partly | n/a | 85.85 | 76.92 | 78.29 | 77.68 |
| COVID-19 policy | Decentralised state-based public health legislation with lockdowns/ social distancing policies; some centralised measures and funding; vaccines available but delayed rollout | Denialism at the federal level; policies implemented locally by governors and majors; moderate lockdown/ local decisions; lack of funding; vaccines applied only after pressure over the President | Strongly decentralised and multi-stakeholder based; moderate to strong lockdown and social distancing policies; public funding to mitigate social effects; vaccines available and easy accessible | Herd immunity prominent throughout; 3 periods of lockdown with Scottish government more conservative than Westminster; public funds for furlough system; vaccines available | National emergency declared early with travel and social restrictions; decentralised policy implementation with high variety; vaccines now available and accessible but strong political controversy |

Sources: authors' own table, based on expert information and the following websites:
\* UNFPA, 2022; https://www.unfpa.org/data/world-population-dashboards
\*\* OECD, 2022; 2021 or latest available data, https://data.oecd.org/gdp/gross-domestic-product-gdp.htm
\*\*\* OECD Health, 2022; latest available data

Our country sample is inspired by more complex qualitative approaches and moves beyond typologies. We consider diversity in relation to the geographical location, the types/institutional conditions of governments and healthcare systems, COVID-19 indicators and pandemic policies, and gender equality measures (Table 1). However, the selected countries must have established (to some degree) gender equality policies, humanitarian rights and democratic political institutions in order to analyse SDG5 targets. Our sample comprises five upper-middle and high-income countries: Australia, Brazil, Germany, the United Kingdom (UK) and United States (USA).

### Collecting and analysing country data comparatively: a qualitative approach

We developed a step-by-step approach, informed by qualitative comparative research. Step 1: Based on the framework shown in Figure 1, a topic guide was developed and agreed among authors, which served as a template for preparing the country cases. Information was collected for each country by the respective author(s) in our team and a first country draft was prepared.

Step 2: The lead authors discussed and reviewed all case studies, identified gaps and queries, harmonised the categories and terminology, and produced a revised, more coherent topic guide. Findings were discussed and adapted by the team; authors revised their country cases accordingly and prepared a more condensed draft.

Step 3: the procedure was repeated until the topic guide was sufficiently coherent and standardised, comprising 19 items related to three major research areas (Table 2). Gaps were closed and the material was summarised in a table prepared for every country (Tables 1.1 to 1.5 Appendix).

**Table 2.**
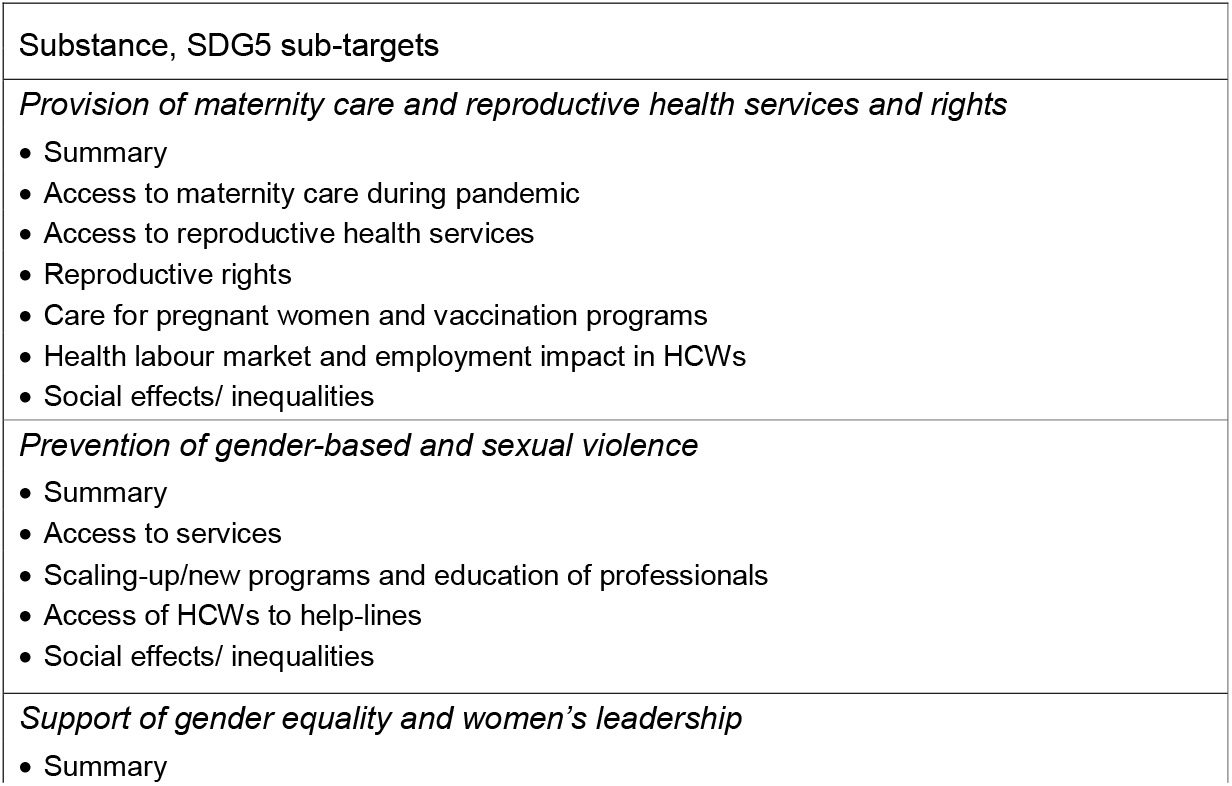
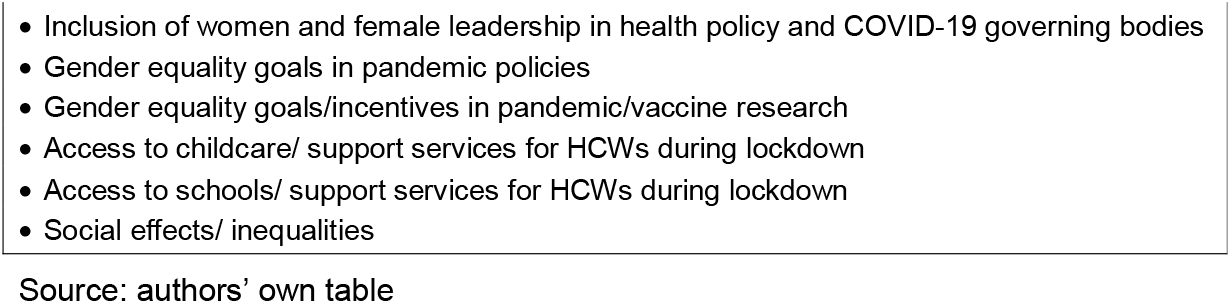
Topic guide for the country case studies

The comparative analysis followed a similar approach. The two lead authors carried out the first analysis across the five cases and prepared a draft manuscript, which all authors discussed and revised. The procedure was repeated until the findings were sufficiently condensed, generalised categories developed and agreement achieved on the findings.

## Results

The three thematic areas (substance) served to structure the comparative analysis. Table 3 summarises the impact of the COVID-19 pandemic in our countries, while Table 4 introduces the action taken and future policy. Some illustrative examples taken from the country cases bring ‘flesh to the bones’ of the tables and provide in-depth information, also paying attention to differences between the countries (see for more detailed references, Appendix Table 1.1-1.5).

**Table 3.** Comparing the impact of COVID-19 in key areas of SDG5 targets in five selected upper-middle and high income countries

| Substance* | Impact of COVID-19 in SDG5 targets, selected countries |
| --- | --- |
| **Maternity care/**<br>**reproductive**<br>**health** |  |
| Summary | Broadly similar trends in maternity care with overall moderate biomedical restrictions but reduced non-medical and social support services; strong limitations in reproductive health services with high regional variation constrain reproductive rights, but relevant differences between countries – US makes the worst case with abortion ban, while Australia and, to a lesser degree the UK took action to protect access to early abortion |
| Maternity care | Essential maternity care services were largely kept open or transformed into digital services; prevention and social support services were significantly limited or suspended |
| Reproductive health services | Strong limitations in all areas of non-essential services, especially through side-effects of lockdown policies and re-allocation of staff due to prioritisation |
|  | of COVID-19 care; relevant differences in policy responses with some support in Australia and UK and worsening in Brazil and the US |
| Reproductive rights | No prioritisation of reproductive health and abortion rights, but little legal restrictions except in the US |
| Care for pregnant women/ vaccination | Some prioritisation of pregnant and breast-feeding women in vaccination programs but often too late |
| Health labour market and employment | Human resources for health were not used effectively; increased COVID19 risk for HCWs; protection of pregnant HCWs varied between countries; HCW shortage exacerbates |
| Social inequalities | Intersecting gender inequalities increased, disadvantaging ethnic minorities, migrants and refugees and more generally vulnerable groups |
| **Gender-based violence** |  |
| Summary | Strong increase in violence but lack of monitoring and reliable data; strong restrictions in access to services; lack of effective violence prevention and protection of women but relevant differences between countries: Australia introduced new programs and increased budgets to scale up support and prevention, while action was poor or lacking in other countries |
| Access to services | Strong restrictions and suspension of services coupled with strong increase in demand and need; digital provision of some services; relevant differences between countries: Australian Government took some action to respond to growing demand, marginal governmental support also in the UK and US, in Brazil and Germany mainly limited to NGOs and municipalities |
| Scaling-up/ new programs and training programs | Overall little attention to quantitative and qualitative changes in demand for services and a need for improving sensitivity; action mainly taken by feminist actors and NGOs, but new governmental support programs and increased budgets in Australia |
| Access of HCWs to help-lines | In some countries 'yes' but overall very little attention to the problem and lack of support with relevant variation between countries |
| Social inequalities | Lack of data but some evidence of reinforced social inequalities with strong intersectional effects, especially connected to digital provision |
| **Gender equality/leadership** |  |
| Summary | Gender equality issues were largely absent from pandemic policies; SDG5 goals and gender mainstreaming policies were ignored; poor participation of women and lack of women's leadership in decision-making; lack of attention to reinforced gender-based disadvantages, especially for mothers |
| Participation/ leadership in pandemic policy | Overall poor participation of women in all areas of pandemic policy and across all levels and actors (except NGOs) involved in governance, including the media and lack of women's leadership |
| Gender equality goals in pandemic policy | Largely absent from pandemic policy and key decisions, like lockdowns; no country applied gender mainstreaming; no prioritisation of gender equality and reproductive rights |
| Gender equality goals in research | Largely absent, no country applied gender mainstreaming guidelines; some mandatory guidelines, especially in vaccine testing; some variation between countries |
| Access to childcare | Strongly restricted; closures during first lockdowns except in Australia, but some availability to HCWs; strong variation in duration of closures and access of HCWs ranging from very little restrictions in Australia to possible loss of childcare in the US |
| Access to schools | Strongly restricted; closures during first lockdowns, but some digital schooling and some on-site services for HCWs; strong variation in access of HCWs and duration of closures |
| Social inequalities | Lack of monitoring but evidence of strong increases; worsening intersecting inequalities |

\* Items are abbreviated (see, Table 2)
Source: authors' own table based on country case reports (Tables 1.1-1.5 Appendix)

**Table 4.** Comparing action taken and future policy plans to protect SDG5 targets in times of COVID-19 pandemic in five selected upper-middle and high income countries

| **Substance\*** | **Action taken and future policy plans to protect SDG5 targets, selected countries** |
| --- | --- |
| **Maternity care/ reproductive health** |  |
| Summary | no country had a policy in place to prioritise and reorganise services; no signs of policy learning and inclusion in future policy; strong expansion of digital services with little attention to inequalities in access; prioritisation of medical care against psycho-social support services; no attention to intersecting inequalities |
| Maternity care | lack of systematic service reorganisation; prioritisation of medical care against psycho-social services; some inclusion of digital services in future policy |
| Reproductive health services | no prioritisation; lack of action and attention, with some exception in Australia |
| Reproductive rights | no prioritisation and policy interventions strongly vary, ranging from some protection and expansion of early medical abortion in Australia (temporarily in the UK) and some improvement (Germany) and no change (Brazil) in legal conditions to a new ban of abortion rights and strong restrictions in some states in the US |
| Care for pregnant women/ vaccination | inclusion in vaccination prioritisation programs but some variation |
| Health labour market and employment | lack of attention and action; some variation especially in relation to protection of pregnant HCWs |
| Social inequalities | no monitoring established; lack of attention and action |
| **Gender-based violence** |  |
| Summary | no prioritisation; lack of prevention and protection of women; action and future policy strongly vary between countries |
| Access to services | action taken varies, ranging from shifting responsibility to NGOs and/or municipalities in Brazil and Germany to significant increases in budgets for support services for victims, new schemes and new laws in Australia, with the UK and US taking position in the middle |
| Scaling-up/ new programs and training programs | new programs in Australia and UK, efforts in other countries focus on filling gaps and ignoring increased demand; lack of training of HCWs and prevention programs with some attention to protection in Australia |
| Access of HCWs to help-lines | little attention |
| Social inequalities | no monitoring established; lack of attention |
| **Gender equality/ leadership** |  |
| Summary | no prioritisation; lack of attention to SDG5 goals and gender mainstreaming; no signs of systematic inclusion in future policy |
| Participation/ leadership in pandemic policy | lack of action; some change and more balanced quotas more recently with new governments in Australia, Germany and the US; some attention from the media |
| Gender equality goals in pandemic policy | lack of attention; no gender mainstreaming |
| Gender equality goals in research | lack of attention; no gender mainstreaming except mandatory requirements in some countries; some action by female scientists in some countries |
| Access to childcare | closures but strong variety in action taken to support HCWs/mothers |
| Access to schools | closures but strong variety in action taken to support HCWs/mothers |
| Social inequalities | no monitoring established; lack of attention |

\* Items abbreviated (see Table 2); Source: authors' own table based on country reports (Appendix)

### Provision of maternity care and reproductive health and rights

All countries tried to keep essential maternity and reproductive services open during the pandemic, but with stronger inclusion of digital services (we refer to ‘digital services’ as an umbrella term including all services provided virtually, e.g. telemedicine, eHealth, mHealth, video/telephone hotlines). The new emergent opportunities of digital provision helped to maintain services. Across countries, we found only incremental change with little direct restrictions in essential maternity care services – looking at access without considering quality and accessibility. The impact was much stronger in non-essential services, where we found severe limitations and suspensions that most strongly affected prevention, counselling and all forms of social support services (e.g. visitors, support of partners during birthing) (Table 3 and Table 4). The situation was more diverse in relation to reproductive health and rights. We observed no change in reproductive rights and little impact on essential services (especially abortion) in some countries but reductions and new abortion bans in others. However, non-essential reproductive health services were significantly restricted in all countries.

Direct policy interventions (such as suspension of services) and lack of attention to women’s healthcare needs may combine and create severe threats that pose risks to women’s lives. In Brazil, 1,088 maternal deaths were documented in 2020/2021 with a weekly average of 22.2 maternal deaths, indicating an increase of 40% in maternal deaths compared to the previous year. Notably, this increase was much higher than in the general Brazilian population where COVID-19 related deaths caused a 29% increase in mortality.

The maternal mortality rate is the most evidence-based indicator of the threats, but it does not show the whole picture (Wenham et al., 2022). In Germany, for instance, we found no signs of higher death rates and overall little signs of increased medical risks (e.g. slightly higher rates of premature birth and caesarean section rates in 2020). However, the strong limitations in support services and preventative care are likely to cause long-term effects on women’s mental health and wellbeing (Schmiedhofer et al., 2022). The Australian case shows that prevalence rates of antenatal depression more than doubled or even nearly tripled compared to pre-pandemic rates; an increase was associated with COVID-19 distress of having a baby during a COVID-19 outbreak (Frankham et al., 2021).

Health workforce policy, in particular the situation of midwives, is another issue for consideration. In the US, poor pandemic policy and high COVID-19 infection rates among midwives caused restrictions of services, which were reinforced through cuts in support services (ante- and post-natal) provided by doulas due to restrictions in visits and accompanying partners. In Germany, new COVID-19 administrative requirements and digital service provision may have reduced services provided by self-employed midwives, who could not afford to pay for additional training and technology. Both cases show that available health workforce capacity is not used effectively to support pregnant women. On the other hand, the case of the UK reveals that pregnant HCWs working in the NHS were poorly protected and exposed to COVID-19 patient care during the first wave; yet legal protection improved and the government introduced mandatory risk assessment for pregnant employees.

Another important issue across countries was the lack of attention and policy focus to the increase in inequalities. Data are overall poor, but our cases indicate that COVID-19 policies may affect vulnerable groups, migrants and asylum seekers more strongly than others (Engelhardt et al., 2021; Lotta, et al., 2021; Wenham et al., 2020). A particular area of concern arises through the replacement of services with digital offerings. For instance, in Brazil, access to virtual maternity care services was more difficult for women who lived in areas without (or with reduced) access to technologies (for international results, see, Galle et al., 2021). Digitalisation may exacerbate inequalities in relation to vulnerable groups, although there may also be some positive effects. No country had adequate policies and programs in place to identify the social effects and mitigate problems.

Regarding reproductive health, we found strong limitations in non-essential services in all countries. Negative effects appear most severe in the US. Women who experienced shifts in their family planning preferences were sometimes not able to obtain contraception or abortion. These developments strongly impacted women’s reproductive rights and have potentially create a policy window for new legislative bans, such as the US Supreme Court decision on abortion (UN Women, 2022). The situation was different in other countries included in our assessment. Lockdown policies affected the availability and accessibility of contraception and prevention services, but did not change abortion rights. In Australia and the UK, access to early medical abortion even improved during the pandemic through inclusion of digital provision. In Germany, physicians’ advertisement of early abortion services was legalised in 2022. This was not directly in response to COVID-19 and other restrictions of abortion and women’ rights remained unchanged.

Despite some important differences in reproductive health and rights, we found strong similarities indicating the need for greater attention. First, no healthcare system had a policy in place to prioritise and reorganise services under COVID-19 conditions and there were little, if any signs of systematic policy learning and prioritisation in future health policy and pandemic recovery plans. Notably, the sustainability of Australian efforts to protect and support early abortion and counselling services in reproductive health is currently not clear. In this situation, changes may happen ‘sideways’ and may be unintended. These intended changes may occur due to limited access during lockdowns, prioritisation of COVID-19 services and shortage of HCWs. Second, pandemic policies fuelled a biomedical approach, while important public health and psycho-social services were significantly weakened or closed due to other priorities. A revival of biomedicalisation (the establishment and dominance of commercialised medical offers for natural phenomena; Christiaens & van Teijlingen, 2009) of maternity care and reproductive health services during COVID-19 happened ‘through the backdoor’ without policy debate and public involvement.

### Prevention of gender-based and sexual violence

Prevention of gender-based and sexual violence was strongly affected by pandemic policy and restrictions. At the same time, need for these services increased dramatically in all countries. We focus on women (and children) who account for the vast majority, but men and non-binary people may also become victims of gender-based violence and in some cases the offender might be female.

Data are overall poor and no country established a monitoring system. Information was mainly drawn from surveys, regional data, or NGO reports. In the UK, for example, two thirds of GPs surveyed before August 2021 said abuse had worsened in the last year due to COVID and exacerbated waiting times. Exact numbers are not available and depend on the data sources, as information from Germany may illustrate (local data, city of Göttingen). According to police data, violence reported to the police offices showed an increase of 9.5% in 2020 and 8.3% in 2021 compared to the previous year. A local feminist emergency service documented a much higher increase: in 2020, 27% more women than in the previous year called for help because of partner violence; 26% more persons called for help because they had observed domestic violence (personal information). In the UK, calls to the Domestic Violence helpline showed a similar figure of a 25% increase. Data are not directly comparable but show a clear trend and are particularly concerning given domestic violence is typically under-reported.

Under-reporting of all forms of sexual violence and discrimination has long been highlighted by feminist organisations. COVID-19 further worsened the situation for several reasons. Women were often not able to call for help because they were living with the offender and lockdowns forced them to stay at home. The COVID-19 disruption also affected the police services and the judicial system that judges cases of violence, as data from the US illustrate: For example, Washington, DC, showed a 43% decrease in patients seeking treatment for sexual assault at one hospital compared to pre-pandemic times; at the same time, women had less access to the legal system and hearings were postponed. Similar problems were reported from Brazil, where the decrease of reported cases was lower (14% in 2020).

Across countries, policies in response to COVID-19 have exacerbated gender-based and sexual violence and limited access to services due to COVID-19 policies. A strong increase in violence (regardless of exact numbers) is a clear indicator of policy failure in all countries. It puts a spotlight on the lack of prevention of violence and protection of women, affecting vulnerable groups the most and exacerbating social inequalities.

There were important differences in the ways governments responded to the problem of domestic violence and the integration of violence prevention into future pandemic plans. In Australia the Federal Government announced the Coronavirus Domestic Violence Support Package in March 2020. This package provided Australian $150 (AUD) million in funding, with AUD 130 million to be provided to state and territory governments to increase frontline family and domestic violence services through a new National Partnership Agreement on COVID-19 Domestic and Family Violence Responses. There was a particular focus on safer housing and emergency accommodation, counselling and outreach, crisis support and helplines, men’s behaviour change programs and other perpetrator interventions, assisting frontline services to manage the demand and explore new technology-based services and delivery methods, and responding to unique challenges in regional, rural, and remote locations (Parliament of Australia, 2020).

Action was also taken by the UK government, which provided 2 million in April 2020 to bolster domestic abuse help lines and online support services. 76 million was pledged in May 2020 to support vulnerable people, of which 25 million was allocated to domestic abuse services. In November 2020, the Ministry of Justice provided a further 10.1 million for rape and domestic abuse support, alongside a further 683,000 from the Home Office. A public awareness campaign was launched by the government to support survivors of domestic abuse. Another important innovation was the establishment of a scheme in partnership with UK pharmacies called ‘Ask for ANI’ (Action Needed Immediately); this code word helped women who could not talk about the offense (e.g. because they live with the offender/could not go out). Lessons drawn from the pandemic also led to the introduction of the Domestic Abuse Act in April 2021, aiming to protect survivors and better address the behaviour of perpetrators.

In contrast, in Brazil and Germany (partly also in the US), action was mainly taken by non-state actors and municipalities. Governments showed very little, if any efforts to step up prevention services and to provide support for those who experienced violence. The findings indicate that NHS systems might provide better opportunities to integrate services related to gender-based violence compared to the other types included in our assessment, but the case of Brazil illustrates that the type of system does not fully predict the policy response in relation to sexual violence.

### Gender equality and women’s leadership

SDG5 targets and women’s leadership were largely absent from the pandemic debate. No country analysed here applied gender mainstreaming and equality policies systematically to organise and govern service provision during the COVID-19 pandemic and there was generally no attention to the monitoring of social inequalities. Across countries, we found limited participation of women in decision-making bodies and a lack of women’s leadership. For instance, in the UK, no gender advisors were included in the Scientific Advisory Group for Emergencies (SAGE) in 2020 and 43% of the daily COVID-19 press conferences featured an all-male line-up with no female politician or expert present. Unsurprisingly, gender was not explicitly considered at any point during the government’s response. For example, in Brazil the government did not establish any national coordination body during the pandemic and most of the legislation about COVID-19 did not include gender issues. In the US, the White House Coronavirus Task Force contained only two female members of 24 total members.

In Germany, we found that women’s voices were marginal in a high-level national think tank. Legally binding guidelines and access of equal opportunity officers to decision-making bodies were ignored when new Corona Task Forces were established at the hospital level. Very few women held leadership positions in relevant academic disciplines; for instance, only 12 percent heads of department in virology are female (Ciesek, 2022). Female experts were also less visible than their male colleagues in the media. Notably, this happened under a female Chancellor during the first 1.5 years of the pandemic. Exacerbating the marginalisation of gender inequality, female experts who raised their voice publicly, often faced hate und strong attacks on social media, and increasingly even physical offences.

We also observed some signs of growing sensitivity to gender inequalities in the media and/or academia. In Germany, for instance, a small NGO ‘ProQuoteMedien’ developed a list of high-level female COVID-19 experts to increase visibility of women in public debate. This action was inspired by the 50/50 program introduced by the BBC in the UK that aimed to achieve a balanced quota of women in the media. Despite some sensitivity – and an increase of women and minorities in government in Australia, Germany and the US – there were little, if any signs of more systematic inclusion of gender equality in future policies. However, it might be too early for recent changes to transform the power structures and governance mechanisms.

The second area of assessment were the programs introduced to mitigate women’s disadvantages caused by gender-based roles and task distribution, with a focus on women with childcare responsibilities. Data from Australia show that women with children reported spending approximately 43 more hours per week on childcare during the pandemic than men. Also, women withdrew from higher education at greater rates than men during the pandemic, with 86,000 fewer women enrolled to study in May 2020 than in May 2019, compared with just over 21,000 fewer men (Zhou, 2020). In all the cases, we found only little effort to provide meaningful support for them. In the UK, the government decided to suspend the mandatory gender pay reporting for employers.

In all countries, childcare facilities and schools faced limitations, especially during the lockdown periods of the first waves of COVID-19 and, in some countries lasting until the end of 2021. These conditions dramatically increased women’s responsibility for childcare and affected the employment and career chances of mothers. Their needs were not systematically included in lockdown policies and schooling schemes, but governments responded in different ways. In the UK, childcare facilities and schools were closed except for children of essential workers, including HCWs. Germany applied a similar policy but with longer periods of closure and variation between states and providers. The situation was worse in the US, where even HCWs sometimes lost childcare support, and in Brazil, where childcare services were closed for 18 months with no support for HCWs. In Australia, in contrast, childcare facilities remained open and the government provided free childcare during the first lockdown. Schools experienced periods of interruption with high variation between states and territories but remained open for essential workers and vulnerable families. The examples highlight that governments may either reinforce or mitigate the disadvantageous effects of COVID-19 in women with childcare responsibilities.

## Discussion

Our comparative assessment reveals similar trends across different health systems and geopolitical and epidemiological contexts. Pandemic policies strongly cut into women’s health and healthcare, but no country has taken action to adequately protect women’s health and rights and to strengthen their voices in the policy process.

The United Nations warned us early, that the COVID-19 pandemic puts the limited gains in gender equality and women’s rights made over the decades at risks of being rolled back (UN Women, 2020c). Available data highlighted the social costs of lockdowns, especially the ‘second pandemic’ (Fiske et al., 2022) and ‘shadow pandemic’: ‘UNFPA had projected that if lockdowns were to continue for 6 months, 31 million additional gender-based violence cases can be expected, and for every 3 months the lockdown continues, an additional 15 million additional cases of gender-based violence are to be expected’ (Feminist COVID Response, Advocacy Monitoring Toolkit, 2021, p. 36). Feminists across the world therefore called to action to protect human rights and the health of women and the UN Secretary General urged ‘governments to put women and girls at the centre of their recovery plans’ (UN, 2020).

However, governments set other priorities. No country prioritised SDG5 goals in the COVID-19 policies and future recovery plans. A global public health emergency response was blind to gender equality and human rights and threatened women’s health and social participation on a large scale (Flor et al., 2022; UNFRA, 2020; UN Women, 2022; Wenham et al., 2020). This holds true for countries with male and female political leaders, for different epidemiological scenarios and lockdown policies, and for various areas of SDG5 and health. This failure raises important questions not only on the SDGs, but also on pandemic policies and recovery plans, including the role of public health institutions (Tomsick et al., 2022).

One explanation of these policy failures might be the creation of a discourse of ‘crisis’ as a global narrative, in which epidemiological measures (daily incidence, contract tracing, personal protective equipment, etc.) and medical and system indicators (death rates, hospital beds, respirators, etc.) dominated public debate and societies. The severity of the global COVID-19 crisis and its disruptions opened policy windows to outflank established democratic institutions and public control. Existing ‘power hubs’ in societies were strengthened, such as politics, science and the media that were better equipped for immediate action to define priorities and transform governance procedures. On an institutional level, men usually dominate these power hubs in terms of numbers and hegemonic interests; new forms of governing provided a power boost because they may act ‘under the radar’ of established gender equality measures (Kuhlmann et al., 2017). In relation to cultural powers, the making of a biomedical discourse of pandemic risk and protection marched in step with reductionist and positivist approaches in science and healthcare (Correia & Willis, 2021). The ‘neutrality’ claims embedded in these approaches made the needs of women, minorities and vulnerable groups invisible and nurtured social inequalities during the pandemic (Fiske et al., 2022; Morgan et al., 202; Wenham et al., 2020). The reasons for exacerbating gender inequalities during the pandemic are therefore complex, yet a global discourse of ‘crisis’ with its own priorities and new powers might explain why we found similar trends across countries with different institutional settings. Discursive governing powers might be strong, regardless of real changes in the institutions, and institutional settings may furnish this discourse with additional power, if met with antifeminist politics. Examples are the ban of abortion rights in the US, or an increase in maternal mortality rates in Brazil. On the other hand, inclusion of reproductive health and violence prevention in public health policy and pandemic responses might counteract these powers to some degree; some efforts were observed in Australia, Germany and the UK (although they were very weak and may not be sustainable). Recent increases in the numbers of women (and minorities) in governments, observed in Australia, Germany and the US, might in future also weaken a gender-blind crisis discourse, although mere changes in sex ratios do not automatically translate into change in gender relations.

### Limitations

A rapid assessment of time-sensitive ongoing developments and policy action in an under-researched area with poor data sources has several limitations. Firstly, data collection was based on secondary sources and some expert information; primary data were mostly unavailable. Secondly, our qualitative comparative approach focused on exploring problems but does not provide representative data. Thirdly, we did not analyse regional differences and our selection of examples may not provide the full country picture. Fourth, we refer to the intersectionality of gender inequality, but a more detailed analysis of complex social inequalities would go beyond the scope of SDG5 and this research. Our comparative study should be read as a pilot that makes gaps in pandemic policies visible and supports feminist calls for action.

## Conclusions

Our assessment highlights a need for gender mainstreaming in COVID-19 policies and recovery plans that should be intersectional to account for the complex social inequalities. A strong connection exists between SDG5 and public health goals. Gender equality, preventative care, mental health and social support services, and the health and wellbeing of HCWs were all weakened by the pandemic and structural inequalities exacerbated, while a reductionist biomedical crisis discourse was revamped. Greater attention to governance may help us to further explore these processes and identify windows of opportunity for feminist actors and policy approaches.

## Supporting information

Appendix country cases summary tables

## Data Availability

All data produced in the present work are contained in the manuscript.

## Acknowledgements

The idea for this study was inspired by a chapter on SDG5 and SDG3 (EK and GL) for the Observatory on Health Systems and Policies/WHO Euro project on co-production of the SDGs (Falkenbach et al., 2022, forthcoming).

## Author contributions

EK and GL had the idea for this study, developed the comparative research framework and wrote a first draft; all authors prepared country case studies (KW, J-LM: Australia; GL, MF: Brazil; EK, LMF: Germany; CW, AH-C: UK; LP: USA), contributed to the comparative analysis and the revisions and have read and approved the final version.

## Data availability statement

Not applicable; relevant data are referenced.

## Supplementary material

A summary table for each country case is provided in the Appendix Tables 1.1-1.5.

## Appendix Tables

Appendix Table 1.1. Australia, country case study summary

Appendix Table 1.2. Brazil, country case study summary

Appendix Table 1.3. Germany, country case study summary

Appendix Table 1.4. United Kingdom, country case study summary

Appendix Table 1.5. USA, country case study summary

