## Appendix country cases summary tables for "SDG5 ‘Gender Equality’ and the COVID-19 pandemic: a rapid assessment of health system responses in selected upper-middle and high-income countries"

### Appendix Table 1.1 Australia, country case study summary

| SDG5 | Impact | Action | Future Policy |
|---|---|---|---|
| **Maternity care/ reproductive health** | | | |
| Summary | essential services were kept open with some limitations and new digital services; restrictions to prevention and support services; evidence of increased rates of antenatal depression; some improvement in reproductive health services and rights | overall little attention but some expansion of reproductive health services, including early abortion; new digital antenatal services are included in Medicare Benefits Schedule (MBS) | some antenatal expansions maintained; currently no decision on reproductive services and rights |
| Maternity care | access to essential services, but restrictions due to pandemic policies (face mask during birthing; no access of partners; etc.) with strong local variation; new digital services and strong expansion; restrictions in support and non-essential services | new digital antenatal services were added to healthcare delivery, covered by the MBS | New digital antenatal services included in the MBS |
| Reproductive services | essential services were kept open but pandemic restrictions on elective surgery included fertility treatments; some variation; early medical abortion was made available via digital consultation; choice of provider for reproductive health consultations | early medical abortion via digital appointments was introduced in the MBS; digital provision of reproductive health consultations was reimbursed by MBS and restriction to choice of provider was relaxed | currently no decision |
| Reproductive rights | some improvement through relaxed legal restrictions of early abortion and through improved MBS rules on choice of provider | no legal action but some improvements through changes in the MBS scheme | currently no decision |
| Care for pregnant women/ vaccination | included in vaccination programs | since June 2021, pregnant women are included in prioritised groups | none |
| Health labour market and employment | higher infection risk of midwives and other HCWs; shortage may worsen | none | none |
| Social inequalities | lack of data; digital services provision may increase social inequality and affect vulnerable groups most strongly; employment loss during the pandemic may restrict fertility decisions | none | none |
| **Gender-based violence** | | | |
| Summary | increase but poor data; some attention to the problem and expansion of services; digital service provision | Government introduced new programs and significantly increased budgets; improved support for victims, some attention to men/offenders and to protection | some attention and new programs |
| Access to services | routine health and legal services were kept open but often moved to digital provision; regional variation; digital services may restrict access | Coronavirus Domestic Violence Support Package introduced in March 2020 including AUS 150 million | some attention and new programs |

| | | | |
|---|---|---|---|
| | | budget and National Partnership Agreement on COVID-19 Domestic and Family Violence Responses; responses to increased demand and new needs, e.g. supporting transformation to digital provision and people in remote/rural areas | |
| Scaling-up/ new programs and training programs | expansion of family violence phone services; expansion of support, e.g. short-term accommodation; improved protective equipment; improved technical support; support for men's behaviour programs; high regional variation; improved awareness but no new training programs to improve sensitivity | significant increase in budgets, including increase in capacity of nationwide family violence services, men's counselling, perpetrator intervention, women's safety at home, support for trafficked people; in Victoria additional funding was made available to meet increased demand | Introduced programs extended or new programs added |
| Access of HCWs to help-lines | some local health services may be available but no national program; high variation | none | none |
| Social inequalities | pandemic restrictions threaten vulnerable groups most strongly; digital provision exacerbates inequalities | lack of attention but some support for people in remote areas to mitigate disadvantages of digital services | none |
| **Gender equality** | | | |
| Summary | lack of attention to gender equality and mainstreaming policy; poor participation of women and female leadership; some support for HCWs with childcare responsibilities | none, except provision of free childcare during the first period of lockdowns | none |
| Women/ female leadership in health policy and COVID-19 governing boards | lack of attention to female leadership and participation in National Advisory Committee; overall lack of data | none; equal opportunity law not systematically applied to pandemic policy and governance; no monitoring | none |
| Gender equality goals in pandemic policy | none | none | none |
| Gender equality goals in research | lack of attention and incentives | none | none |
| Access to childcare | facilities were kept open with a few exemptions; regional variation | Government provided free childcare during lockdown in 2020; in case of closures services were kept open for HCWs and vulnerable families | none |
| Access to schools | periods of closures but strong regional variation; move to digital schooling | schools were kept open for essential workers/HCWs and vulnerable families | none |
| Social inequalities | increase in gendered inequalities with intersectional effects | lack of attention but some support for remote areas | none |

Source: authors' own table, based on expert information, published secondary sources

Key references

Australian College of Midwives (ACM) (2020). Women's experiences of maternity care at the height of COVID-19 ACT: Australian College of Midwives. https://www.midwives.org.au/news/womens-experiences-maternity-care-height-covid-19

Boxall, H., Morgan, A., & Brown, R. (2020). The prevalence of domestic violence among women during the COVID-19 Pandemic. Statistical Bulletin no. 28. Australian Institute of Criminology. https://www.aic.gov.au/publications/sb/sb28

Department of Health and Human Services (2020). Visiting hospitals: rest of Victoria Melbourne. Australia Department of Health and Human Services. https://www.dhhs.vic.gov.au/visiting-hospitals-covid-192020

Frankham, L.J., Thorsteinsson, E.B., & Bartik, W. (2021). Antenatal depression and the experiences of Australian women in the maternity system during the COVID-19 pandemic. Open Journal of Depression, 10, 155–167. https://www.scirp.org/journal/ojd

Johnston, R.M., Mohammed, A., & van der Linden, C. (2020). Evidence of exacerbated gender inequality in child care obligations in Canada and Australia during the COVID-19 pandemic. Politics & Gender, 16(4),1131–1141. doi:10.1017/S1743923X20000574

National Health and Medical Research Council (2021). National COVID-19 Health and Research Advisory. NHMRC. https://www.nhmrc.gov.au/about-us/leadership-and-governance/committees/national-covid-19-health-and-research-advisory-committee

Parliament of Australia (2020). Family violence in Australia and the National Plan. Parliament of Australia. https://www.aph.gov.au/Parliamentary_Business/Committees/House/Social_Policy_and_Legal_Affairs/Familyviolence/Report/section?id=committees%2Freportrep%2F024577%2F75208

Pfitzner, N., Fitz-Gibbon, K., & True, J. (2020). Responding to the 'shadow pandemic': practitioner views on the nature of and responses to violence against women in Victoria, Australia during the COVID-19 restrictions. Monash University, Victoria, Australia. https://apo.org.au/sites/default/files/resource-files/2020-06/apo-nid306064.pdf

Royal Australia and New Zealand College of Obstetricians and Gynaecologists (RANZCOG) (2020). COVID-19 Hub Melbourne. RANZCOG. https://ranzcog.edu.au/statements-guidelines/covid-19-statement

Swannell, C. (2020). Early medical abortion: telehealth restrictions discriminatory. The Medical Journal of Australia, 21, 1. https://www.mja.com.au/system/files/2020-09/FINAL%2021%20SEP%20EMA.pdf

Wood, D., Griffiths, K., & Crowley, T. (2021). Women's work: the impact of the COVID crisis on Australian women. Grattan Institute. https://grattan.edu.au/report/womens-work/

Zhou, N. (2020). Female enrolment at Australian universities dropped by 86,000 in 2020 as 'pink recession' hit. The Guardian. https://www.theguardian.com/australia-news/2020/nov/12/female-enrolment-at-australian-universities-dropped-by-86000-in-2020-as-pink-recession-hit

### Appendix Table 1.2 Brazil, country case study summary

| SDG5 | Impact | Action | Future Policy |
|---|---|---|---|
| **Maternity care/ reproductive health** | | | |
| Summary | essential services were kept open but with limited access; strong restrictions to prevention and support services; new digital services; 40% increase in maternal deaths in 2020/21 compared to previous year | none; a general trend of decreasing healthcare budgets since 2015 exacerbated the impact of COVID-19 | maintenance of essential maternity care services during health crises |
| Maternity care | limited access to/ accessibility of services due to pandemic restrictions and re-allocation of HCWs; new digital service delivery | none except new digital services; service restrictions due to COVID-19 prioritisation | maintenance of primary and specialised services during health crises |
| Reproductive services | limited access to services due to pandemic restrictions and move of HCWs; new digital services | none, except new digital services; services reduced due to COVID-19 prioritisation | none |
| Reproductive rights | no public debate; no explicit restrictions | none; support of maintenance of legal abortion by feminist initiatives, including moving service to digital offers | none |
| Care for pregnant women/ vaccination | inclusion in vaccination programs, but too late | initial prioritisation of all pregnant women suspended by Ministry of Health in May 2021 and restricted to proof of co-morbidities; criteria of the Operational Plan for comorbidities were very strict and did not cover all situations of gestational risk | none |
| Health labour market and employment | strong shortage of HCWs in primary and specialised care regardless due to prioritisation of COVID-19 services; higher infection risk of HCWs providing COVID-19 care | none | none |
| Social effects/ inequalities | increase in social inequalities; maternal death rate was higher among low-income and black women; new digital services may exacerbate inequalities | none; efforts of civil society groups to raise awareness | none |
| **Gender-based violence** | | | |
| Summary | increase of vulnerability but underreporting; reduced opportunity to report; closure of public defenders' offices and transformation to digital services; decrease of 14% in denounces in 2020; strong regional variety; lack of data | intention to create a National Plan against Femicide (Dec. 2020) by federal Government but no signs of implementation; closure of specialised services; reduced collection of data; efforts by municipalities, NGOs and private actors to improve sensitivity and support services | none |

| | | | |
|---|---|---|---|
| Access to services | women's policy stations remained open but limited opportunity to report violence; strong restrictions in access to all services; new digital services; digital provision reduced access of women who lived with the aggressor; services strongly decentralised with high variety and regional differences. | none on national level; efforts by some municipalities, NGOs and private companies to bypass national policy, to improve reporting (e.g. digital) and provide services; support by global NGOs | none |
| Scaling-up/ new programs and training programs | strong limitations due to pandemic priorities and restrictions; strong regional variation; some transformation to digital services | none on national level; some efforts by municipalities, NGOs and private companies to mitigate the problem, keep services open and introduce new programs, including digital offers; some support by global NGOs (e.g. UNICEF) | none |
| Access of HCWs to help-lines | limited access and longer waiting times; increased need; 30% of HCWs surveyed 2020-21 reported abuse had worsened | none on national level; some municipalities introduced mental health services for HCWs | none |
| Social effects/ inequalities | increased vulnerability of women living with the aggressor; digitalisation exacerbates social inequalities; lack of data | none | none |
| **Gender equality** | | | |
| Summary | participation of women and female leadership were very poor and weakened; strong political dominance of men; lack of data | none; gender equal goals and monitoring were absent; male dominance in government was strengthened; a National Plan against Feminicide was initiated but not implemented; | none |
| Women/ female leadership in health policy and COVID-19 governing boards | no; Bolsonaro government has one of the lowest rates in the history of women ministers; all high-level positions related to the pandemic are held by men | none; all high-level positions related to the pandemic were provided to men; replacements in the Health Ministry were filled by men only; equal opportunity goals not applied to pandemic policy and governance; no monitoring established; | none |
| Gender equality goals in pandemic policy | None; no prioritisation of all pregnant women in the vaccination calendar, but some municipalities changed the status to priority after strong complaints | none; general lack of pandemic policies and establishment of specific decision-making bodies on national level; some attention on the level of municipalities | none |
| Gender equality goals in research | None | none, lack of attention and data | none |
| Access to childcare | closed during 2020/21 and reopened in November 2021 | none | none |
| Access to schools | closed during 2020/21 with online classes and fully reopened in November 2021 | none | none |
| Social effects/ inequalities | increase in intersecting inequalities as women live more often in vulnerable conditions; lack of data | none | none |

Source: authors’ own table, based on expert information and published secondary sources

### Key references

Amorim, M.M.R., Takemoto, M.L.S., & Fonseca, E.B.D. (2020). Maternal deaths with coronavirus disease 2019: a different outcome from low- to middle-resource countries? American Journal of Obstetrics & Gynecology, 223(2), 298–299. https://pubmed.ncbi.nlm.nih.gov/32348744/

Diniz D., Brito, L., & Rondon, G. (2022). Maternal mortality and the lack of women-centered care in Brazil during COVID-19: preliminary findings of a qualitative study. Lancet Regional Health – Americas,10, 100239. https://www.thelancet.com/journals/lanam/article/PIIS2667-193X(22)00056-4/fulltext

Ferigato, S., Fernandez, M., Amorim, M., Ambrogi, I., Fernandes, L.M.M., & Pacheco, R. (2020). The Brazilian Government's mistakes in responding to the COVID-19 pandemic. Lancet, 396(10263), 1636. https://pubmed.ncbi.nlm.nih.gov/33096042/

Fernandez, M., & Amorim, M.M.R. (2021). Morte de grávidas e puérperas por COVID-19. Rede Brasileira de Mulheres Cientístas. Nota Técnica, 2021, 1. https://mulherescientistas.org/wp-content/uploads/2021/05/Nota-Tecnica-n.1-Gravidas-e-Puerperas.pdf

Fórum Brasileiro de Segurança Pública. Violência doméstica durante a pandemia de Covid-19. (2020, April 16). Forum Brasileiro de Segurança Pública. https://forumseguranca.org.br/wp-content/uploads/2020/06/violencia-domestica-covid-19-ed02-v5.pdf

Jaccoud, L., Sátyro, N., Gomes, S., Vieira, F., Servo, L., & Fernandez, M. (2021). Por que a coordenação nacional de políticas públicas importa para os direitos dos cidadãos, especialmente na pandemia? Rede Brasileira de Mulheres Cientístas. Nota Técnica, 2021, 11. https://mulherescientistas.org/wp-content/uploads/2021/07/NT-11.pdf

Marques, E.S., Moraes, C.L., Hasselman, M.H., Deslandes, S.F., & Reichenheim, M.E.A. (2020). Violência contra mulheres, crianças e adolescentes em tempos de pandemia pela COVID-19: panorama, motivações e formas de enfrentamento. Cad. Saúde Pública, 36(4), e00074420. https://doi.org/10.1590/0102-311X00074420

Menezes, M.O., Takemoto, M.L.S., Nakamura-Pereira, M., Katz, L., Amorim, M.M.R., Salgado, H.O., Melo, A., Diniz, C.S.G., de Sousa, L.A.R., Magalhães, C.G., Knobel, R., & Andreucci, C.B. (2020). Brazilian Group of Studies for COVID-19. Risk factors for adverse outcomes among pregnant and postpartum women with acute respiratory distress syndrome due to COVID-19. International Journal of Gynaecology and Obstetrics, 151(3), 415–423. doi: 10.1002/ijgo.13407

Silva, V.R., & Ferreira, L. (2020, July 2). Só 55% dos hos dos hospitais que ofereciam serviço de aborto legal no Brasil seguem atendendo na pandemia. Gênero e Número. http://www.generonumero.media/so-55-dos-hospitais-que-ofereciam-servico-de-aborto-legal-no-brasil-seguem-atendendo-na-pandemia/

Takemoto, M.L.S., Menezes, M.O., Andreucci, C.B., Knobel, R., Sousa, L.A.R., Katz L., Fonseca, E.B., Magalhães, C.G., Oliveira, W.K., Rezende-Filho, J., Melo, A.S.O., & Amorim, M.M.R. (2020). Maternal mortality and COVID-19. Journal of Maternal-Fetal & Neonatal Medicine, 16, 1–7. doi: 10.1080/14767058.2020.1786056

Wenham, C., Fernandez, M., Corrêa, M.G., Lotta, G., Schall, B., Rocha, M.C., & Pimenta, D.N.G (2021). Gender and race on the frontline: experiences of health workers in Brazil during the COVID-19 pandemic. Social Politics: International Studies in Gender, State & Society, online. doi:10.1093/sp/jxab031

Zignoni, C. (2020, April 9). O governo que odeia as mulheres: a inércia de Damares Alves na crise do Covid-19. INESC. https://www.inesc.org.br/o-governo-que-odeia-as-mulheres-a-inercia-de-damares-alves-na-crise-da-codiv-19/

### Appendix Table 1.3 Germany, country case study summary

| SDG5 | Impact | Action | Future Policy |
|---|---|---|---|
| **Maternity care/ reproductive health** | | | |
| Summary | essential services were kept open; restrictions to prevention and support services; new digital services; some evidence of slightly higher rates of premature births and cesarean deliveries; lack of comprehensive epidemiological data | action was limited to vaccination recommendation and expansion of digital services; in 2022 legal change to de-criminalise physicians' advertisement of early abortion | no attention beyond routine services and existing mandatory legal requirements |
| Maternity care | access to essential services, but pandemic restrictions (e.g. face mask during birthing; no access of partners; delayed admission in case of COVID-19 infection); local variation; strong restrictions of non-essential services, especially prevention and counselling; digital service provision | new digital services; reimbursement of digital services through social health insurance funds | implementation of digital services in routine care under consideration |
| Reproductive services | essential services remained open but pandemic restrictions; suspension/ limitations in prevention and support services; staff moved to COVID-19 services; regional/ organisational variation; digital provision | none, except transformation of some services to digital provision | none |
| Reproductive rights | no restrictions, but negative impact through limited access and availability of services | no change to rights/restrictions, but decriminalisation of physicians' advertisement of early abortion services | none |
| Care for pregnant women/ vaccination | included in vaccination programs but delayed recommendation by national authorities, also for breast-feeding women | none, except mandatory inclusion of women in vaccine studies; no midwifery experts in pandemic policy | none |
| Health labour market and employment | high infection risk of midwives; digital services more difficult for self-employed midwives due to high costs and training needs; may reinforce HCW shortage due to drop-outs | none, except mandatory infection prevention law to exclude pregnant midwives and other HCWs from patient care, with full compensation | none |
| Social inequalities | increased inequalities; increased discrimination; restricted prevention and support services threaten all vulnerable groups (migrants, asylum seekers, ethnic minorities, elderly) most strongly; digitalisation exacerbates inequalities | none; some local variation may apply; some action taken by feminist and migrant networks/ NGOs; no systematic data and monitoring available | none |
| **Gender-based violence** | | | |
| Summary | some attention to the problem; data vary between sources but all show strong increases in all forms of violence; e.g. increase > 100%, during quarantine 7.5% of women/ | none; some action taken by NGOs and feminist networks; some media attention but not connected to pandemic policy | none |

| | | | |
|---|---|---|---|
| | 10.5% of children experienced sexual violence | | |
| Access to services | routine health and legal services were kept open but prevention services strongly limited due to lockdown and allocation of staff to pandemic prevention; increase in digital services; NGOs/ feminist groups are important service providers but with limited resources | no response to new needs; lack of coherent monitoring/ data are scattered and not standardised; feminist activists not included in pandemic governance | none |
| Scaling-up/ new programs and training programs | feminist advocacy and some service expansion but no new approaches/ training programs; restrictions due to pandemic policy; regional variation | None | none |
| Access of HCWs to help-lines | routine services and occupational health services for hospital staff; no specific help-lines for HCWs | none; no support from health professional associations, but some student groups set up online platforms | none |
| Social inequalities | limited access to services and pandemic policies threaten vulnerable groups most strongly; new need for targeted services for migrants/asylum seekers, minorities; digitalisation worsens inequalities | none | none |
| **Gender equality** | | | |
| Summary | participation of women and female leadership poorly developed and weakened; male scientists more visible in the media; lack of data | none, gender mainstreaming and equal opportunity policy were largely ignored; some sensitivity and more balanced gender composition in new Coalition Government | none, except some general gender equality targets |
| Women/ female leadership in health policy and COVID-19 governing boards | weak female leadership and participation; no spill-over from female Chancellor (until autumn 2021); women and female groups poorly represented in pandemic policy; no mandatory inclusion of equal opportunity officers in hospital COVID-19 Task Forces; weak media presence of female scientists | none; equal opportunity law not systematically applied to pandemic policy and governance; lack of transparency; no monitoring system; more recently some attention to female COVID-19 scientists | none |
| Gender equality goals in pandemic policy | largely ignored on all levels | none | none |
| Gender equality goals in research | lack of data and incentives; little public attention | none, except mandatory inclusion of female participants in vaccine testing | none |
| Access to childcare | for essential workers/ HCWs, but often strongly limited and not reliable due to staff shortage | none | none |
| Access to schools | closed during the first wave; re-open with some restrictions/ uncertainties | none, but critical public debate, including attention to increased workload of women | none |
| Social inequalities | increase in gendered inequalities with strong intersectional effects | lack of political attention | none |

Source: authors' own table, based on expert information and secondary sources

Key references

Allmendinger, J. (2020). Kritik am Leopoldina Statement: Das Wohlergehen der Frauen wird nicht adressiert. Tagesspiegel. https://www.tagesspiegel.de/wissen/kritik-an-leopoldina-empfehlung-das-wohlergehen-der-frauen-wird-nicht-adressiert/25739444.html

Bariola, N., & Collins, C. (2021). The gendered politics of pandemic relief: labor and family policies in Denmark, Germany, and the United States during COVID-19. American Behavioral Scientist. 65(12), 1671–169. https://journals.sagepub.com/doi/pdf/10.1177/00027642211003140

Ciesek, S. (2022). Virologie im Medienfokus: Lehren aus der Corona-Krise. Labourjournal, Juli/August, 8-11. https://www.laborjournal.de/rubric/essays/essays2022/e22_01.php

Czymara, C.S., Langenkamp, A., & Cano, T. (2021). Cause for concerns: gender inequality in experiencing the COVID-19 lockdown in Germany. European Societies, 23 (Suppl1), S68–81. https://www.tandfonline.com/doi/full/10.1080/14616696.2020.1808692

Engelhardt, M., Krautstengel, A., Patzelt, L., Gaudion, M., Kamhiye, J., & Borde, T. (2021). Auswirkungen der Covid-19 Pandemie auf die Versorgungssituation von geflüchteten Frauen während Schwangerschaft und Geburt. Zeitschrift für Geburtshilfe und Neonatologie, 225(S 01), P141. https://doi.org/10.1055/S-0041-1739903

Pro Familia Medizin (2020). Corona Krise: Reproductive Health Facts – ein Zwischenstand. Nr 1, 1–16. https://www.profamilia.de/fileadmin/dateien/fachpersonal/familienplanungsrundbrief/pro_familia_medizin_1-2020.pdf

Hagenbeck, C., Pecks, U., Fehm, T., Borgmeier, F., Schleußner, E., & Zöllkau, J. (2020). Pregnancy, birth, and puerperium with SARS-CoV-2 and COVID-19. Gynäkologe, 53(9), 614–623. https://doi.org/10.1007/S00129-020-04637-9/TABLES/2

Hipp, L., & Bünning, M. (2021). Parenthood as a driver of increased gender inequality during COVID-19? Exploratory evidence from Germany. European Societies, 23(S1), S658–S673. https://doi.org/10.1080/14616696.2020.1833229/SUPPL_FILE/REUS_A_1833229_SM7875.DOCX; https://www.tandfonline.com/doi/pdf/10.1080/14616696.2020.1833229?needAccess=true

Schmiedhofer, M., Derksen, C., Dietl, J.E., Häussler, F., Louwen, F., Hüner, B., Reister, F., Strametz, R., & Lippke, S. (2022). Birthing under the condition of the COVID-19 pandemic in Germany: interviews with mothers, partners, and obstetric health care workers. Int. J. Environ. Res. Public Health, 19(3), 1486. https://doi.org/10.3390/ijerph19031486

Steiner, J., & Ebert, C. (2020). Tatort Wohnzimmer: Gewalt gegen Frauen und Kinder im Corona Lockdown. Ärztetag Podcast. https://www.aerztezeitung.de/Podcasts/Tatort-Wohnzimmer-Gewalt-gegen-Kinder-und-Frauen-im-Corona-Lockdown-410287.html

Zoch, G., Bächmann, A.-C., & Vicari, B. (2020). Care arrangements and parental well-being during the COVID-19 pandemic in Germany. LIfBi Working Paper No. 91. Leibniz Institute for Educational Trajectories. https://doi.org/10.5157/LIfBi:WP91:2.0

### Appendix Table 1.5 United Kingdom, country case study summary

| SDG5 | Impact | Action | Future Policy |
|---|---|---|---|
| **Maternity care/ reproductive health** | | | |
| Summary | essential services remained open but limited access and some replacement by digital provision; strong limitations in non-essential and support services; increase in social inequalities | maternity care was defined as essential service but restrictions due to pandemic policies and lack of resources; no prioritisation of reproductive health; some action by NHS England to support mothers from minority groups; variation due to devolution | none |
| Maternity care | essential services remained open but access was limited; strong limitations in support service; limited partner support; new digital services | maternity care was defined as essential service but lack of attention and resources | none |
| Reproductive services | strong limitations in provision and access to services due to pandemic restrictions and closure of services; new digital services | no prioritisation of services; introduction of new digital services in NHS services; in March 2020 temporary support of abortion services through availability of early abortion pills without medical supervision | none |
| Reproductive rights | temporary legal change | temporary legalisation (March 2020) of at-home administration of early abortion pills, previously only available for second pill | permanent availability of at-home administration in England/ Wales; Scotland to decide |
| Care for pregnant women/ vaccination | inclusion in vaccination programs | none, except vaccination recommendation | none |
| Health labour market and employment | higher infection risk of HCWs providing COVID-19 care; poor protection and poor worker rights of pregnant HCWs | lack of full legal protection; risk assessment of pregnant women by employer introduced | none |
| Social inequalities | increase in social inequalities, especially affecting minority and vulnerable groups | none; some action by NHS England to support pregnant women from minority groups | none |
| **Gender-based violence** | | | |
| Summary | strong increase of sexual violence; some expansion of services and digital provision, but limited access due to pandemic restrictions | increased budgets/public funding of services; new law in 2021 to better protect survivors; introduction of digital services | none |
| Access to services | services were kept open or provided digital, but limitations due to COVID-19 restrictions | increased budgets and public funding of services; improved financial support for NGOs/Charities providing services; digital service | none |

| | | | |
|---|---|---|---|
| | | provision; expansion of access through inclusion of pharmacists | |
| Scaling-up/ new programs and training programs | some expansion of services; some new programs; inclusion of new professional groups; no new training schemes | expansion of providers; new emergency program 'Ask for ANI' (Action Needed Immediately) in collaboration with pharmacists; launch of public awareness campaign; Domestic Abuse Act 2021 aims to better protect survivors and address the behaviour of perpetrators | none |
| Access of HCWs to help-lines | no specific program | none | none |
| Social inequalities | lack of data, but increased disadvantage of minorities and vulnerable groups, especially through digital services | none | none |
| **Gender equality** | | | |
| Summary | participation of women was limited and female leadership lacking; lack of attention to gender equality goals; increase in intersecting inequalities | none | Scotland to embed the UN CEDAW convention into Scots Law |
| Women/ female leadership in health policy and COVID-19 governing boards | marginalisation of women in high-level media events; in 2020 43% of daily COVID-19 press conferences featured only male politicians and experts; participation in the Scientific Advisory Group of Experts (SAGE) ranged between 33%-44% and was nearly balanced 2021, but no gender advisor was included | none; equal opportunity goals were not applied to pandemic policy | none |
| Gender equality goals in pandemic policy | lack of attention | none, except inclusion of COVID-19 vaccination in pregnancy monitoring and attention to men's higher risk of severe COVID-19 disease; worsening legal conditions and data through suspension of gender pay gap reporting for employers | none |
| Gender equality goals in research | none | none | none |
| Access to childcare | facilities were closed during lockdowns; open for essential workers including HCWs | none | none |
| Access to schools | Schools were closed during first lockdown; open for essential workers including HCWs; remained open for all during second lockdown | none | none |
| Social inequalities | increase in intersecting inequalities affecting ethnic minorities and vulnerable groups most strongly; digital service provision worsens inequalities; career disadvantages | none | none |

| | |
|---|---|
| | of women with childcare responsibilities |

Source: authors' own table, based on expert information and secondary sources

Key references

Bradbury-Jones, C., & Isham, L. (2020). The pandemic paradox: the consequences of COVID-19 on domestic violence. Journal of Clinical Nursing, 29, 2047–2049. https://onlinelibrary.wiley.com/doi/epdf/10.1111/jocn.15296

Herten-Crabb, A., & Wenham, C. (2022). "I was facilitating everybody else's life. And mine had just ground to a halt": the COVID-19 pandemic and its impact on women in the United Kingdom. Social Politics: International Studies in Gender, State & Society, online. https://doi.org/10.1093/sp/jxac006

Iacobucci, G. (2021). Covid-19: Female NHS and care staff report deteriorating health because of pandemic. BMJ, 373, n1157. https://doi.org/10.1136/bmj.n1157

Jardine, J., Relph, S., Magee, L., von Dadelszen, P., Morris, E., Ross-Davie, M., Draycott, T., & Khalil, A. (2021). Maternity services in the UK during the coronavirus disease 2019 pandemic: a national survey of modifications to standard care. BJOG: An International Journal of Obstetrics & Gynaecology, 128(5), 880–889. https://doi.org/10.1111/1471-0528.16547

Karavadra, B., Stockl, A., Prosser-Snelling, E., Simpson, P., & Morris, E. (2020). Women's perceptions of COVID-19 and their healthcare experiences: a qualitative thematic analysis of a national survey of pregnant women in the United Kingdom. BMC Pregnancy and Childbirth, 20(1), 600. https://doi.org/10.1186/s12884-020-03283-2

Kourti, A., Stavridou, A., Panagouli, E., Psaltopoulou, T., Spiliopoulou, C., Tsolia, M., Sergentanis, T. N., & Tsitsika, A. (2021). Domestic violence during the COVID-19 pandemic: a systematic review. Trauma, Violence, & Abuse, online. https://doi.org/10.1177/15248380211038690

Mansour, D. (2021). Maintaining sexual and reproductive health services in the UK during COVID-19. BMJ Sexual & Reproductive Health, 47(4), 235–237. https://doi.org/10.1136/bmjsrh-2021-201142

Quach, G. (2022, January 21). Only two UK Covid briefings were led by a female MP, report finds. The Guardian. https://www.theguardian.com/world/2022/jan/21/only-two-uk-covid-briefings-female-mp-2022-sex-and-power-index

Strauss, C., & Patel-Campbell, C. (2021). May 2021 update: COVID-19 and the female health and care workforce survey. NHS Confederation. https://www.nhsconfed.org/publications/may-2021-update-covid-19-and-female-health-and-care-workforce-survey

UK Government. (2021, January 14). Pharmacies launch codeword scheme to offer 'lifeline' to domestic abuse victims. GOV.UK. https://www.gov.uk/government/news/pharmacies-launch-codeword-scheme-to-offer-lifeline-to-domestic-abuse-victims

UK Women's Budget Group. (2021, March 31). One year on: women are less likely than men to feel the Government's response to Covid-19 has met their needs. https://wbg.org.uk/analysis/reports/one-year-on-women-are-less-likely-than-men-to-feel-the-governments-response-to-covid-19-has-met-their-needs/

### Appendix Table 1.5 USA, country case study summary

| SDG5 | Impact | Action | Future Policy |
|---|---|---|---|
| **Maternity care/ reproductive health** | | | |
| Summary | decrease of prenatal care visits; increase in demand for mental healthcare; increase in maternal mortality rate (23.8 per 100,000 live births); access to maternity care services was significantly limited but partly replaced by digital services | new digital services included in COVID-19 emergency policies, but strong variation | extension of emergency digital maternity care and some permanent provision, but high variation |
| Maternity care | significantly limited access to services and reduced accessibility due to a general decrease and pandemic restrictions; COVID-19 restrictions strongly weaken social/ partner support and support of doulas; new digital service delivery | significant cuts in services due to COVID-19 policy; new digital services introduced in most insurance programs (e.g. audio- calls combined with monitoring and health coaching); high variation | extension of emergency digital maternity care and some permanent provision; high variation |
| Reproductive services | limited access to services due to pandemic restrictions and closure of services; new digital services | no prioritisation of services or piecemeal implementation in state-led emergency responses; new digital services but lack of infrastructures | Department of Health and Human Services established Reproductive Healthcare Access Task Force in 2022 |
| Reproductive rights | strong limitations or even suspension of abortion rights through both limited access to services and new legal decisions/ suspension of Roe vs Wade; high variation between states | 14 states suspended abortion, only 12 states explicitly protected abortion; medication abortion facilitated by telemedicine is prohibited in 18 states; some attempts to deny Medicaid coverage for certain methods of contraception; high variation between states | suspension of abortion through High Court decision but efforts to protect abortion rights |
| Care for pregnant women/ vaccination | inclusion in vaccination programs, but lack of attention to the needs of COVID-19 positive women | tailored messaging to pregnant and breast-feeding women to promote vaccination | none |
| Health labour market and employment | higher infection risk of HCWs providing COVID-19 care; some task-shifting to doulas to mitigate cuts in maternity care provision | none | none |
| Social inequalities | increase in social inequalities; disruption of reproductive services hit vulnerable populations most strongly; restrictions of Medicaid programs most strongly affected low income people; delays or cancellation of reproductive services were higher in Black and LGBTQIA populations; digital services disadvantaged vulnerable group | none; digital services were thought to reduce racial disparities but the overall effects are less clear and may reinforce inequalities | none |
| **Gender-based violence** | | | |

| | | | |
|---|---|---|---|
| Summary | strong increase of sexual violence while access to services was significantly limited; some underreporting due to closed services; early in the pandemic minors made up half of visitors to national sexual assault hotline | none | none |
| Access to services | strong limitations; one-third of women reported difficulty accessing resources after violence incidents (2020); reduced access to legal services due to courts being closed | some digital service provision | none |
| Scaling-up/ new programs and training programs | some new programs; some exemption from closures during lockdown; strong regional variation; some transformation to digital services; no new training programs | 21 states enacted protections for violence survivors; 5 states included explicit exemptions from non-essential business closures for providers; feminist NGOs increased support services | none |
| Access of HCWs to help-lines | no information on specific programs | none | none |
| Social inequalities | inequalities exacerbated for under-served and vulnerable populations; lack of data | none | none |
| **Gender equality** | | | |
| Summary | participation of women and female leadership were overall poor; some recent increase in numbers; lack of attention to gender equality goals; lack of data | none | none |
| Women/ female leadership in health policy and COVID-19 governing boards | no signs of high-level female leadership but quotas may be higher at lower levels; women accounted for 10% in White House Coronavirus Task Force, but for 84% of staff of Centres for Disease Prevention and Control; data are overall poor | none; equal opportunity goals were not applied to pandemic policy; some increase in female and minority participation in the new Government (2022) | none |
| Gender equality goals in pandemic policy | lack of attention | none | none |
| Gender equality goals in research | none | none; a criticism of COVID-19 emergency policies | none |
| Access to childcare | strong restrictions and some closures; some HCWs lost childcare support affecting women most strongly; high variation | none | none |
| Access to schools | schools were closed; after re-opening were only small numbers of students accepted; some exceptional opening for HCWs but strong variation | none | none |
| Social inequalities | increase in gender inequalities most strongly in vulnerable populations; digitalisation worsens inequalities | none | none |

Source: authors' own table, based on expert information and secondary sources

Key references

Adams C. (2022). Pregnancy and birth in the United States during the COVID-19 pandemic: The views of doulas. Birth, 49(1), 116–122. https://doi.org/10.1111/birt.12580

Ahlers-Schmidt, C.R., Hervey, A.M., Neil, T., Kuhlmann, S., & Kuhlmann, Z. (2020). Concerns of women regarding pregnancy and childbirth during the COVID-19 pandemic. Patient Education and Counseling, 103(12), 2578–2582 https://doi.org/10.1016/j.pec.2020.09.031

Aiken A.R.A., Starling, J.E.,Gomperts, R., Tec, M., Scott, J.G., & Aiken, C.E. (2020) Demand for self-managed online telemedicine abortion in the United States during the Coronavirus disease 2019 (COVID-19) pandemic. Obstetrics and Gynecology, 136(4), 835–837. https://doi.org/10.1097/AOG.0000000000004081

Connor, J., Madhavan, S., Mokashi, M., Amanuel, H., Johnson, N.R., Pace, L.E., & Bartz, D. (2020). Health risks and outcomes that disproportionately affect women during the Covid-19 pandemic: a review. Social Science & Medicine, 266, 113364. https://doi.org/10.1016/j.socscimed.2020.113364

Flor, L. S., Friedman, J., Spencer, C.N., Cagney, J., Arrieta, A., Herbert, M.E., Stein, C., Mullany, E., Hon, J., Patwardhan, V., Barber, R.M., Collins, J.K., Hay, S.I., Lim, S.S., Lozano, R., Mokdad, A.H., Murray, C.J.L., Reiner, R.C. Jr., Sorensen, R.J.D., Pigott, D. M., Haakenstad, A., & Gakidou, E. (2022). Quantifying the effects of the COVID-19 pandemic on gender equality on health, social, and economic indicators: a comprehensive review of data from March, 2020, to September, 2021.The Lancet, 399, P2381–2397. https://doi.org/10.1016/S0140-6736(22)00008-3

Gutschow, K., & Davis-Floyd, R. (2021). The impacts of COVID-19 on US maternity care practices: a follow-up study. Frontiers in Sociology, 6, 655401. https://doi.org/10.3389/fsoc.2021.655401

Maier, M., Samari G., Ostrowski, J., Bencomo, C., & McGovern, T. (2021). 'Scrambling to figure out what to do': a mixed method analysis of COVID-19's impact on sexual and reproductive health and rights in the United States. BMJ Sexual & Reproductive Health, 47(4), e16. http://dx.doi.org/10.1136/bmjsrh-2021-201081

Robinson, L.J., Engelson, B.J., & Hayes, S.N. (2021). Who is caring for health care workers' families amid COVID-19? Academic Medicine: Journal of the Association of American Medical Colleges, 96(9), 1254–1258. https://doi.org/10.1097/ACM.0000000000004022

Sapire, R., Ostrowski, J., Maier, M., Samari, G., Bencomo, C., & McGovern, T. (2022). COVID-19 and gender-based violence service provision in the United States. PLoS ONE, 17(2), e0263970. https://doi.org/10.1371/journal.pone.0263970

Stratton, P., Gorodetsky, E., & Clayton, J. (2021) Pregnant in the United States in the COVID-19 pandemic: a collision of crises we cannot ignore. Journal of the National Medical Association, 2021, 113(5), 499–503. https://doi.org/10.1016/j.jnma.2021.03.008

Voth Schrag, R.J., Leat, S., Backes, B, Childress, S., & Wood, L. (2022). 'So many extra safety layers': virtual service provision and implementing social distancing in interpersonal violence service agencies during COVID-19. Journal of Family Violence, online. https://doi.org/10.1007/s10896-021-00350-w
